# Mental disorders and adherence to antiretroviral treatment in health facilities in Mozambique

**DOI:** 10.1101/2022.07.17.22277384

**Authors:** Flavio M. Mandlate, M. Claire Greene, Luis F. Pereira, Maria Lidia Gouveia, Jair Jesus Mari, Francine Cournos, Cristiane S. Duarte, Maria A. Oquendo, Marcelo Feijó Mello, Milton L. Wainberg

**Author notes:** Corresponding author: Flavio M. Mandlate, Ministerio da Saude (Ministry of Health), Eduardo Mondlane Avenue, nr 1008, Maputo City, Postal Code 264, Mozambique. Senior co-authors (Marcelo Feijó Mello, Milton L. Wainberg). E-mail addresses of authors: Flavio M. Mandlate M. Claire Greene Luis F. Pereira Maria Lidia Gouveia Jair Jesus Mari Francine Cournos Cristiane S Duarte Maria A. Oquendo Milton Wainberg Marcelo Feijó Mello.

## Abstract

**Introduction:** Less adherence to antiretroviral treatment (ART) has been found among people suffering from HIV (PWH) with comorbid mental disorders like depression and alcohol use in Mozambique, a Sub-Saharan African country. However, less is explored with regards to other mental disorders.

**Methods:** This study assessed the association of multiple mental disorders and adherence to ART based on the data from primary/tertiary health care facilities in Maputo and Nampula, Mozambique. We administered a sociodemographic questionnaire, Mini International Neuropsychiatric Interview (MINI) Plus 4.0.0 adapted for use in Mozambique to assess mental conditions, and a 3-item self-report to measure ART adherence.

**Results:** Out of 1469 participants, 409 were HIV positive (self-report), with an average age of 36.7 years (SD=9.8), and 30.4% were male. The most common mental disorders were major depressive disorder (27.34%) followed by psychosis (22.03%), suicidal ideation/behavior (15.44%), and alcohol-use disorder (8.35%). Higher levels of non-adherence to ART [(Mean Difference=1.19, 95% CI: 1.04, 1.37)] and the likelihood of missing at least one dose in the last 30 days (OR=3.06, 95% CI: 2.00, 4.67) were found in participants with any mental disorder compared to those without a mental disorder. The highest levels of non-adherence were observed among those with drug use disorders and panic disorder.

**Conclusions:** In Mozambique, PWH with any co-occurring mental conditions had a lower probability of ART adherence. Integrating comprehensive mental health assessment and treatment and ART adherence interventions tailored to PWH with co-occurring mental disorders is necessary to attain optimal ART adherence and reach the UNAIDS ART target.

## Introduction

The introduction of effective antiretroviral therapy (ART) in 1996 significantly changed the scenario of HIV epidemic in high-income countries (1). Unfortunately, the global effort to include low-income countries in this treatment drive did not succeeded until 2001 (2, 3). Initially, when ART was introduced, 95% adherence to ART was required to achieve maximal viral suppression (4). Consistent adherence to ART with consequent undetectable HIV viral load increases the life expectancy of people with HIV (PWH) (5) and prevents new HIV infections (6). The current UNAIDS 95-95-95 global effort to end the HIV epidemic by 2030 emphasizes that 95% of people with HIV know their status, 95% of those knowing their status have access to treatment, and 95% of those under treatment have suppressed viral load (7). Factors associated with poor adherence to ART in sub-Saharan countries include unavailability of health care services, stigma, and discrimination, mental health problems (e.g., hazardous alcohol use, depression), negligence to take medications, poor understanding of HIV illness, disbelief about HIV and its treatment, poverty, food insecurity, medication side effects, poor patient-provider relationships, and difficulty accessing health services (8, 9, 10, 11, 12, 13, 14, 15, 16, 17, 18).

Studies in sub-Saharan Africa and other low-income countries have reported that patients with depression, anxiety, and PTSD symptoms during enrollment in HIV care have shown delayed initiation of ART and negligence to care (19). Moreover, an association of anxiety, depression, and alcohol use has been found with poor adherence to HIV-treatment (20, 21, 22, 23, 24), which in turn leads to high mortality and other poor HIV-related outcomes (25, 26, 27, 28, 23, 24, 29, 30, 31, 32).

Studies evaluating the prevalence of mental disorders among PWH in Sub-Saharan Africa, the world’s region with the highest prevalence of HIV/AIDS, have reported the high prevalence of depression, hazardous alcohol use, post-traumatic stress disorder, anxiety disorders, psychosis, bipolar disorders, and suicidal ideation (33, 26, 34, 35, 36, 37, 38; 39) among HIV patients. However, data regarding the association of poor adherence to ART and comorbid mental disorders besides depression and hazardous alcohol use among PWH, is inconsistent. Some studies from high-income countries have found no association (4, 40, 41) while others reported that depression, anxiety, bipolar disorder, substance use disorder, and suicidal behavior are associated with poor adherence (42, 27, 28). Noticeably, anxiety was strongly associated with poor adherence to treatment among PWH in six studies from Asia while four from sub-Saharan Africa did not find any association (43). Among patients with HIV and co-morbidities such as bipolar disorder, nonadherence to psychiatric medication may lead to ART non-adherence with poor outcomes for both conditions (44). The assessment of adherence to ART among bipolar patients is difficult to measure depending on the tool used (44, 45). However, few studies have also shown the contradictory data. Wagner et al in Los Angeles (USA), found high rates of adherence to ART among patients with both serious mental illness and HIV infection (46). These findings are not in line with those from low-income countries. This could be because of poor knowledge of illness, lack of recognition of medication benefits, and negligence to take medication. Adrawa et al also showed an association between alcohol use and non-adherence in community-based ART distribution settings in Uganda, with participants attributing forgetfulness as a major cause of nonadherence (47). Available data suggest that addressing mental health problems by early screening and interventions at the time of counseling, testing, ART initiation, and follow-up care may be critical for better outcomes for PWH (48, 49, 50).

Mozambique is among countries with highest prevalence rates of HIV viz. 13.2% in 2015 and 12.6% in 2018 among individuals between 15 to 49 years (51, 52). Although various studies have been conducted in Mozambique which reported rates of adherence to ART (53, 54), to the best of our knowledge none have reported association between non-adherence to ART and mental health problems.

To promote adherence to ART treatment in low-resource settings, routine screening and treatment for alcohol use disorder (55) and depression (56) are recommended (26; 34). Unfortunately, lay health workers and healthcare professionals who provide HIV/AIDS treatment services often lack the training and skills to identify and treat co-occurring mental disorders in PWH (29). In Mozambique, similar to most of the low- and middle-income countries (LMICs), screening and treatment for mental conditions have not been integrated into HIV care yet. Psychosocial counselors supervised by psychologists are trained to offer ART adherence counseling but not to identify or treat mental disorders (57).

Keeping all this in view, in this study we examined demographic and clinical correlates of adherence to ART among PWH attending two primary health care facilities and one general hospital in the Maputo City metropolitan region, and three health centers in Nampula City. Poor adherence to ART was hypothesized among PWH with mental disorders than those without a co-morbid mental illness. To the best of our knowledge, this is the first study to assess, if any association exists between mental disorders in PWH and adherence to antiretroviral treatments in Mozambique.

## Methods

### Participants and procedures

Adults from 6 primary and tertiary health care facilities in Mozambique who reported being HIV+ (n=409) were included in this study. Participants were recruited from 2 primary healthcare facilities (health centers) and 1 tertiary facility in Maputo (general hospital) between May 16 and Jun 8, 2018, and from 3 primary healthcare facilities in Nampula between Nov 28 to Dec 13, 2018. These participants were selected from 1469 adults who had been recruited into a cross-sectional validation study conducted to develop a screening tool for multiple mental disorders (58). Patients and those accompanying them were considered eligible for the study, if they were 18 years of age or older, did not display or reported any cognitive impairment, and provided written consent to participate in the study. Participants were assessed in Portuguese using culturally adapted instruments administered by trained research assistants, including demographic questions and a diagnostic interview for mental disorders. All questionnaires and tools used in the study have been either previously validated or translated and adapted using the WHO recommended method (59). Data were collected with tablets using the REDCap electronic data management system. Participants provided written informed consent as approved by the New York State Psychiatric Institute Institutional Review Board (#7479) and the Ethics Council of Eduardo Mondlane University (CIBS FM & HCM/54/2017).

### Measures

*Demographic questions* included information regarding age, sex, the reason for attending the hospital (patient, accompanier, or others), marital status, educational level, occupation, and household income. We also recorded information about the study site (Maputo or Nampula) and facility type (primary vs. tertiary care).

*Mental Disorders (MD)* were assessed using a previously published structured diagnostic interview, the Brazilian version of the Portuguese language Mini-International Neuropsychiatric Interview (MINI) Plus 4.0.0 adapted for use in Mozambique (60, 61). The following modules were administered and included in the analysis: major depressive disorder, dysthymic disorder, panic disorder, generalized anxiety disorder, post-traumatic stress disorder, somatization disorders, mania, psychotic disorders, alcohol use disorder, drug use disorder, and suicidal instinct.

*HIV status and antiretroviral treatment adherence* were self-reported. All participants reported whether they had HIV in the assessment for chronic diseases done in the interview. Participants who were HIV-positive were queried for ART. Participants on ART were further questioned about the duration of ART and how frequently they go to a medical unit to obtain ART medication. Furthermore, they were asked the following three questions about ART adherence from a 3-item self-report measure for medication adherence, which together forms an *ART adherence composite index* ranging from zero to 100, with scores less than 100 indicating some non-adherence (62):

1. In the last 30 days, how many days did you miss at least one dose of ART medication?
2. In the last 30 days, how well did you stick to taking the ART medication in the way you were supposed to? Response options: Very poor, Poor, Reasonable, Good, Very Good, Excellent
3. In the last 30 days, how often did you take your HIV medicine in the way you were supposed to? Response options: Never, Rarely, Sometimes, Habitually, Almost always, Always

As recommended in previous research, the ART adherence composite index was calculated by linearly transforming responses for the three adherence items to a 0-100 scale with zero representing worst adherence and 100 indicating best adherence (62).

### Statistical Analysis

Bivariate associations were calculated between ART adherence and 1) socio-demographics and 2) mental disorders using unadjusted logistic regression models among participants who were HIV+ and responded to the questions about HIV ART adherence (N=395). To control the potential confounders of the association between mental disorders and ART adherence, we reported the adjusted odds ratio and 95% confidence interval controlling for sociodemographic characteristics that were significantly associated with individual indicators of ART adherence (sex, site). Logistic regression model was used to establish the association between any non-adherence (*binary variable*) and specific mental disorders and categories of disorders (common mental disorder, severe mental disorder, moderate/severe suicide risk, and substance use disorder). Among participants who reported any non-adherence based on the composite adherence index, we examined the association between specific mental disorders and categories with the level of non-adherence (*continuous variable ranging from 0-100)* in this subgroup. The reference group for each of these models was PWH participants without any mental disorder comorbidities.

## Results

Of the 409 participants who self-reported to be HIV-positive, the analytic sample was restricted to HIV positive participants on ART (n=395, 96.58%). Among these 395 participants, 44.81% met the criteria for at least one mental disorder. The most common mental disorders were: major depressive disorder (27.34%) followed by psychotic disorders (22.03%), any suicidal ideation (15.44%), alcohol use disorder (8.35%), mania (6.84%), generalized anxiety disorder (6.08%), post-traumatic stress disorder (4.05%), panic disorder (3.29%), drug use disorder (1.27%), somatization disorders (0.76%), and dysthymic disorder (0.51%). Most of the PWH participants were recruited from Maputo (73.92%) and in primary healthcare settings (70.89%). The mean age of participants was 36.7 years (SD=9.8). Majority (69.6%) of the participants were females. More than half of the participants were married (58.0%). Most of the participants had less than secondary school education (80.8%), and a mix of occupations and income levels which have been represented in Table 1. Relative to participants who reported complete adherence to ART in the past 30 days, those that were not fully adherent were less likely to be females (OR=0.53, 95% CI: 0.31, 0.90) and less likely to be recruited from Nampula (OR=0.23, 95% CI: 0.14, 0.38).

### ART adherence and mental disorder

Most of the participants who were on ART reported taking these medications for more than two years (71.1%) and going to the health center monthly to pick up their medication (60.0%), followed by every three months (30.9%) and irregularly (9.1%) (Table 2). Almost three-quarters of the patients (74.68%) reported some non-adherence on at least one of the three questions that comprised the ART composite adherence index in the past 30 days. The presence of a mental disorder was not found to be associated with how long participants had been on ART or how often they went to the health centers to pick up ART. In the unadjusted models, a significant number of participants with any mental disorder reported non-adherence (79.66%) defined as any missed doses, less than excellent adherence to medication regimen, or not always taking medication as prescribed relative to those without a mental disorder (70.64%; OR=1.62, 95% CI: 1.02, 2.60). When adjusting for sex and site, the association between having a mental disorder and any non-adherence was no longer found to be significant (OR=1.57, 95% CI: 0.96, 2.56).

We also assessed the association between ART non-adherence and specific types of mental conditions controlling for site and sex (Table 3). Severe mental disorders, including mania and psychotic disorders, displayed increased odds of non-adherence relative to people without mental disorders. Other mental disorders also displayed a similar trend, but the difference was not statistically significant. We do not report the parameter estimates for dysthymia, somatization, and drug use disorders, given the small number of participants meeting these disorders’ criteria and the limited variation in adherence.

Association between ART adherence and specific types of mental disorders was also evaluated. Individuals with any severe mental disorder (OR=2.38, 95% CI: 1.24, 4.56), mania (OR=4.66, 95% CI: 1.03, 21.27), or psychotic disorder (OR=2.33, 95% CI: 1.19, 4.56) displayed greater odds of non-adherence relative to individuals without a mental disorder. Among the 295 participants without perfect adherence, we found lower adherence among those with any mental disorder (Mean Diff=-2.99; 95% CI: -5.80, -0.18), panic disorder (Mean Diff=-13.79, 95% CI: -20.98, -6.60), generalized anxiety disorder (Mean Diff=-4.95, 95%

CI: -9.64, -0.27), any severe mental disorder (Mean Diff=-3.87, 95% CI: -7.01, -0.73), psychotic disorder (Mean Diff=-4.39, 95% CI: -7.69, -1.09), and any suicide risk (mean Diff=-4.25, 95% CI: -8.13, -0.37) relative to individuals without any mental disorder (Table 3; Figures 1a-b).

**Figure 1a.**
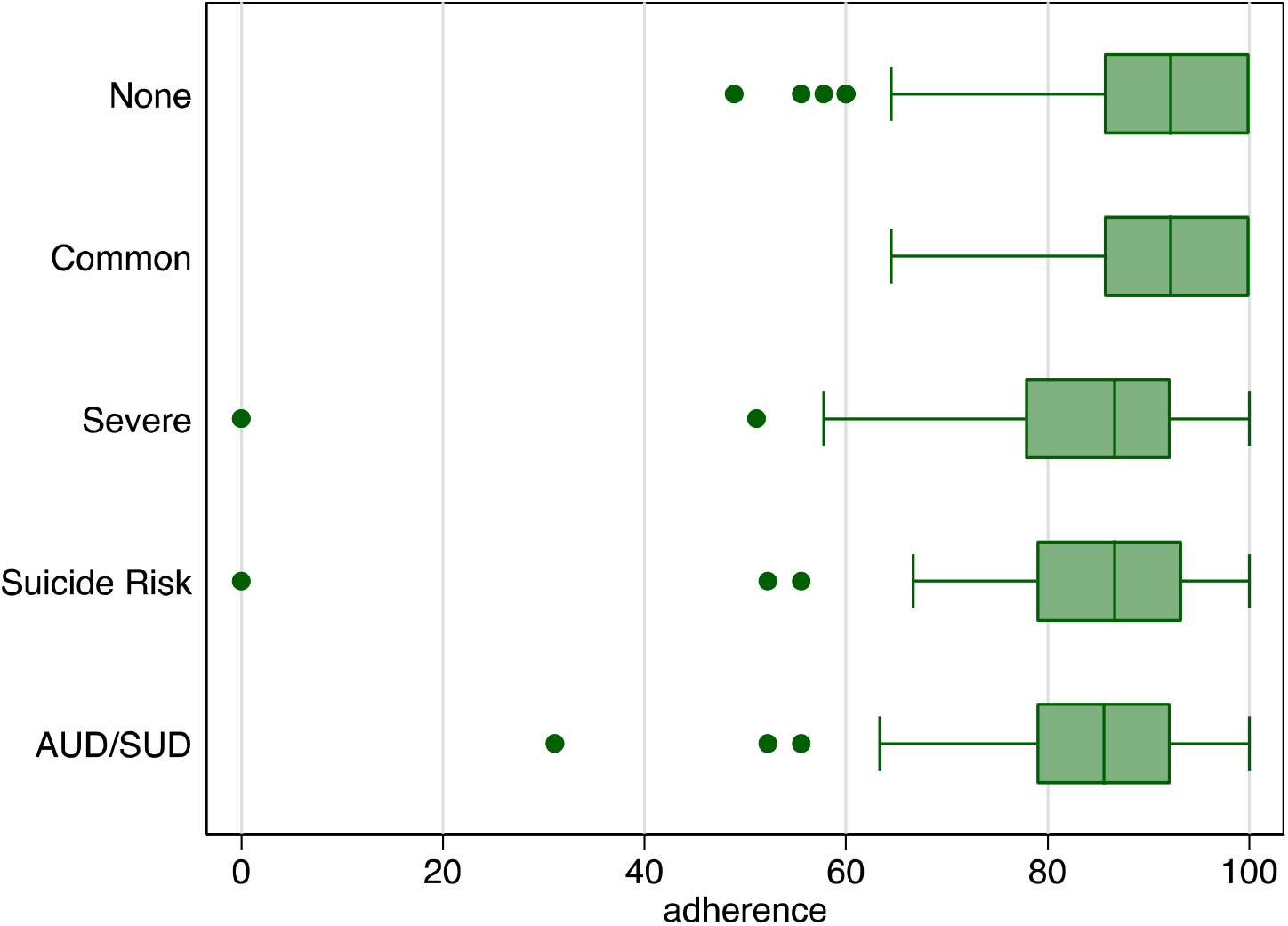
Distribution in level of medication adherence by mental disorder categories.

**Figure 1b.**
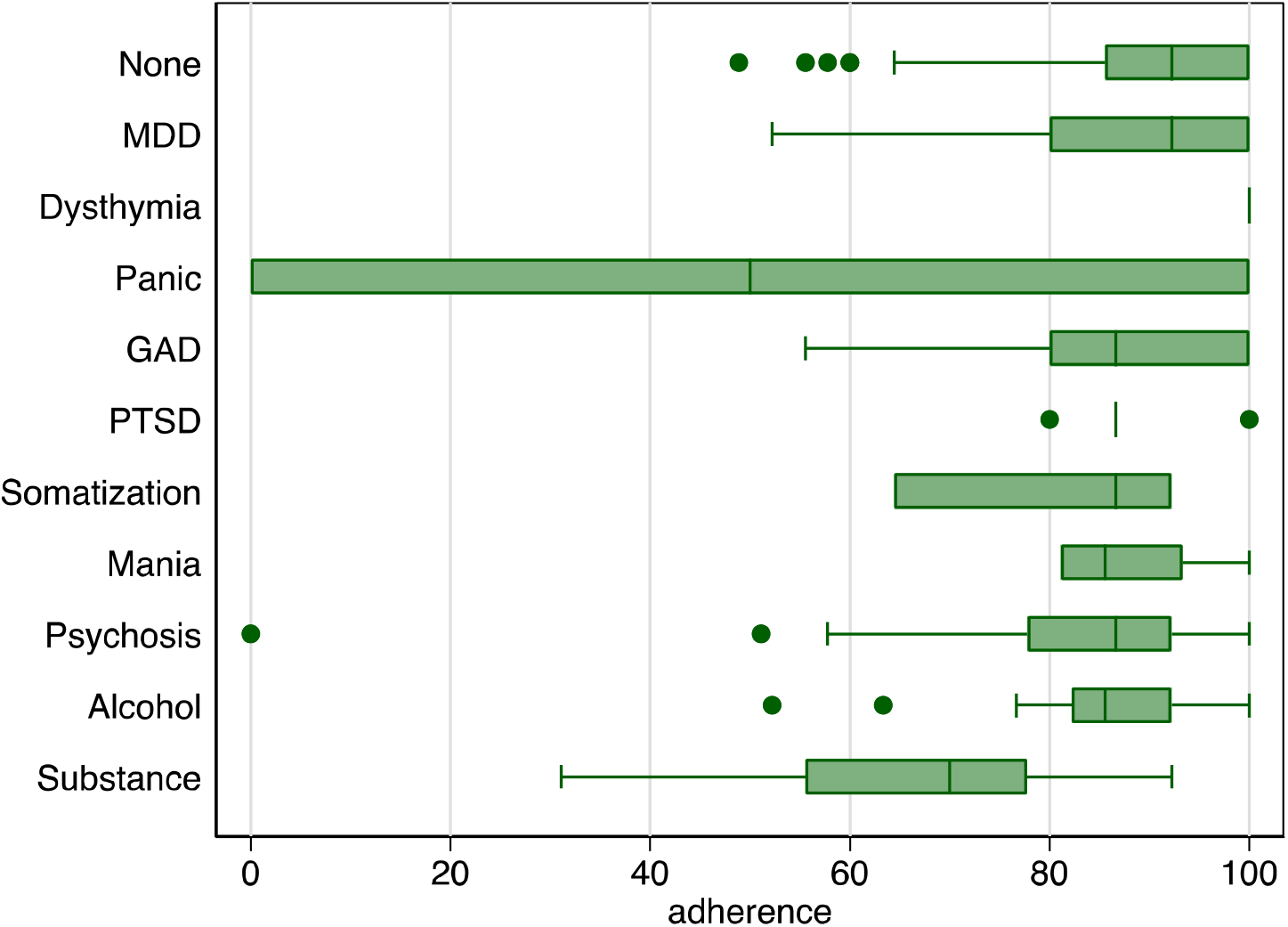
Distribution in level of medication adherence by mental disorder diagnoses.

## Discussion

Our study found that among PWH who were taking ART, almost half (44.81%) of the individuals met the criteria for at least one mental disorder. This finding is consistent with other studies where between 33.4%-50% of PWH were reported to have any comorbid mental disorder (63, 64).

Most participants in our sample (75%) reported non-adherence to ART in the last 30 days (see Tab.1), which is consistent with a previous study in Mozambique reporting that 77-83% of PWH were non-adherent to ART (65). As in previous studies, we found that younger age was marginally associated with non-adherence to ART (29, 66, 67).

In our study, only severe mental illness, which included mania and psychosis, was associated with low adherence. This association could be explained by poor reality testing, insight and judgment, impulsiveness, and disorganization of behavior and thought process, which may impair patients’ ability to take a medication daily (68, 69). However, as suggested by Bogart et al, the use of specialized programs that include access to care management and mental health treatment for patients with severe mental illness may help to narrow the health disparities in this population. They found that following these measures for PWH, there was no difference in the use of ART among patients with severe mental illness when compared with the group without mental illness (70).

In the sub-analysis of participants who did not report complete adherence, several other mental disorders were associated with increased odds of non-adherence. Although four studies from sub-Saharan Africa did not find an association between anxiety and poor adherence in PWH, six studies from Asia did describe a strong association, which is similar to our findings (43). It is possible that taking ART may serve as a reminder of having a chronic and stigmatized illness, with a subsequent increase in anxiety.

We did find any association between drug use disorders and poor adherence. On the contrary to previous studies in sub-Saharan Africa, we did not identify any association between alcohol use disorder and ART adherence (71; 72). Given that the level of non-adherence among participants with alcohol use disorder in our sample were among the highest, we anticipated that this null finding may be explained, in part, by the analyses being underpowered.

An association between depression and poor adherence has been frequently described in the literature. However, in our sample, MDD was not significantly associated with decreased adherence, like a study from Brazil (73). Of note, our findings showed some discrepancies in the association of poor adherence to ART and comorbid mental disorder, mainly regarding depression with other published data (74; 75; 76; 77; 78), but noticeably, there was an association of depressive symptoms with non-adherence within the past (one) month, although not in the past week. This was in line with a previous study which also did not find any associations between mood and suicidality symptoms and missed doses in the past week or month (77).

As in our study, two cross-sectional studies did not find a statistically significant association between schizophrenia/psychotic disorder and ART adherence, using a self-report measure ACTG (Adult AIDS Clinical Trials Group adherence questionnaire) 3 - days recall and EDMs (electronic drug monitors) to evaluate adherence to ART (80).

The Mozambique Ministry of Health has been implementing task-shifting in their mental health system of care since 1996 by training mid-level mental health specialists supervised by psychiatrists or psychologists in district-level clinics (81; 82). Almost 300 psychiatric technicians have been trained to provide mental health care in primary healthcare facilities throughout urban areas of the country. But 70% of the population lives in rural areas, with limited coverage by mental health professionals, with only one psychiatric technician available per district. It’s therefore challenging to scale up comprehensive mental health services integrated into primary care in rural areas (82, 83).

This study has some limitations. First, this is a cross-sectional study, and biological markers did not confirm HIV status and ART adherence. We cannot make causal inferences about the direction of the observed relationship between ART adherence and mental conditions, nor are we able to rule out sources of information bias (e.g., reporting bias and recall bias which can arise in self-report questionnaires) and misclassification. Second, we did not obtain information about what specific ART regimens participants were taking and specific adherence to each ART prescribed. Treatment regimens may have affected the results as several antiretrovirals - including efavirenz, which is still commonly used in Mozambique - cause neuropsychiatric side effects, including depression, anxiety, suicidal ideation, psychosis, and mania. This study did not capture all potential confounders of the association between mental conditions and ART adherence, including CD4 levels and ART regimen (66). Third, this study recruited a convenience sample of PWH in primary and tertiary healthcare facilities and does not represent Mozambique’s general population. The validation study from where our data was obtained was designed to recruit participants with various mental disorders within primary care, HIV care, and mental health clinics. Thus, our findings regarding the proportion of people with mental disorders should not be considered prevalence rates. Fourth, the number of participants meeting the criteria for some mental disorders in our sample was low, resulting in unstable estimates for some conditions (e.g., drug use disorder, somatization disorders). Given the small sample sizes in subgroups, many of the models may have produced unstable and imprecise estimates. Thus, these analyses should be interpreted as exploratory. Future research with larger sample sizes is needed to evaluate the relationship between these low-prevalence mental conditions and ART adherence.

Despite these limitations, this study has several strengths, including using a fully structured diagnostic interview to diagnose most mental disorders resulting in a sample with a breadth of mental conditions and replicating a composite index assessing ART adherence instead of a single-item indicator. Finally, to our knowledge, this is the first study to examine the relationship between ART adherence and its negative association with multiple mental disorders in Mozambique and sub-Saharan Africa, a region with a high burden of HIV/AIDS.

## Conclusions

We found that PWH with co-occurring mental disorders were less likely to be optimally adherent to ART in Mozambique and therefore less likely to accomplish the 95-95-95 targets. These findings support the integration of comprehensive mental health screening in HIV care in Mozambique, together with ART adherence interventions tailored to PWH with comorbid mental conditions. Future research should study methods to expand current psychosocial support interventions beyond the more common approaches targeting depression and alcohol use disorders. Our study shows the importance of identifying and treating most mental disorders among PWH as they also negatively impact ART adherence, quality of life, and the 95-95-95 targets. Researchers should consider using valid ART adherence measures and interventions and rigorous, and validated comprehensive mental health assessments (58) with accompanying evidence-based treatments (83) to be implemented through task-shifting approaches integrated within HIV care in Mozambique and throughout the region.

## Data Availability

All data produced in the present work are contained in the manuscript

## Authors contributions

FM, MCG, LFP drafted the manuscript; MCG did statistical analysis; MLG, JJM, CSD, FC, MFM, MLW reviewed the manuscript; CSD, MAO, MLW designed the protocol. All authors agreed to the final version of the manuscript.

## Authors information

MLW and MFM are both co-senior authors

## Acknowledgments

We thank the Mozambican Ministry of Health, National Directorate of Public Health, Maputo City Health Department, Nampula Province Health Department, and research assistants for facilitating data collection and all study participants for their contribution to this work.

## List of abbreviations

ACTG: Adult AIDS Clinical Trials Group adherence questionnaire
AIDS: Acquired Immunodeficiency Syndrome
ART: Antiretroviral Treatment
ARV: Antiretroviral
AUD: Alcohol Disorder
AUDIT: Alcohol Use Disorder Tool
ART: Highly Active Antiretroviral Therapy
EDM: electronic drug monitor
HIV/AIDS: Human Immunodeficiency Virus/ Acquired Immunodeficiency Syndrome
MDD: Major Depressive Disorder
MD: Mental Disorder
MH: Mental Health
MINI: Plus Mini-International Neuropsychiatric Interview Plus 4.0.0
PWH: People Living With Human Immunodeficiency Virus/ Acquired Immunodeficiency Syndrome sSA – Sub-Saharan Africa

