## Supplementary material for "Mental disorders and adherence to antiretroviral treatment in health facilities in Mozambique": Tables

**Table 1: Sociodemographic correlates of ART adherence among HIV positive participants on ART (n=395)**

|  | **On ART 395 (100.00%)** | **Complete adherence**  **(past 30-days)**  **100 (25.32%)** | **Any non-adherence (past 30-days)**  **295 (74.68%)** | **OR** | **95% CI** |
| --- | --- | --- | --- | --- | --- |
| Age (yrs); mean (sd) | 36.66 (9.82) | 38.24 (8.81) | 36.12 (10.10) | 0.98 | 0.95, 1.00 |
| % Female; n (%) | 275 (69.62) | 79 (79.00) | 196 (66.44) | 0.52 | **0.31, 0.90** |
| Site, n (%) |  |  |  |  |  |
| Maputo | 292 (73.92) | 51 (51.00) | 241 (81.69) | REF | REF |
| Nampula | 103 (26.08) | 49 (49.00) | 54 (18.31) | 0.23 | **0.14, 0.38** |
| Health facility, n (%) |  |  |  |  |  |
| Primary healthcare | 28­0 (70.89) | 72 (72.00) | 208 (70.51) | REF | REF |
| Tertiary healthcare | 115 (29.11) | 28 (28.00) | 97 (29.49) | 1.08 | 0.65, 1.78 |
| Marital status, n (%) |  |  |  |  |  |
| Married/Civil Union | 229 (57.97) | 59 (59.00) | 170 (57.63) | REF | REF |
| Single | 129 (32.66) | 29 (29.00) | 100 (33.90) | 1.20 | 0.72, 1.99 |
| Widowed, divorced, separated | 37 (9.37) | 12 (12.00) | 25 (8.47) | 0.72 | 0.24, 1.53 |
| Education level, n (%) |  |  |  |  |  |
| Less than primary | 92 (23.29) | 24 (24.00) | 68 (23.05) | REF | REF |
| Less than secondary/technical | 227 (57.47) | 58 (58.00) | 169 (57.29) | 1.03 | 0.59, 1.79 |
| Secondary/technical | 67 (16.96) | 15 (15.00) | 52 (17.63) | 1.22 | 0.58, 2.56 |
| University or above | 9 (2.28) | 3 (3.00) | 6 (2.03) | 0.71 | 0.16, 3.05 |
| Occupation, n (%) |  |  |  |  |  |
| Unemployed | 108 (27.34) | 30 (30.00) | 78 (26.44) | REF | REF |
| Formal job | 145 (36.71) | 42 (42.00) | 103 (34.92) | 0.94 | 0.54, 1.64 |
| Informal job | 103 (26.08) | 17 (17.00) | 86 (29.15) | 1.95 | 0.99, 3.80 |
| Full-time student | 12 (3.04) | 1 (1.00) | 11 (3.73) | 4.23 | 0.52, 34.20 |
| Other | 27 (6.84) | 10 (10.00) | 17 (5.76) | 0.65 | 0.27, 1.59 |
| Income (Mzn), n (%) |  |  |  |  |  |
| 0 to 2.800 (0-1 minimum salaries) | 45 (11.39) | 16 (16.00) | 29 (9.83) | REF | REF |
| 2801-5000 (1-2 minimum salaries) | 124 (31.39) | 33 (33.00) | 91 (30.85) | 1.52 | 0.73, 3.15 |
| 5001-15,000 (2-3 minimum salaries) | 90 (22.78) | 22 (22.00) | 68 (23.05) | 1.71 | 0.78, 3.71 |
| >15,000 (3 or more minimum salaries) | 20 (5.06) | 3 (3.00) | 17 (5.76) | 3.13 | 0.79, 12.31 |
| Missing | 116 (29.37) | 26 (26.00) | 90 (30.51) | 1.91 | 0.90, 4.04 |

**Table 2: Association between ART adherence and mental disorder among PWH on ART (n=395)**

|  | **No mental disorder comorbidities**  **218 (55.19%)** | **Mental disorder comorbidities**  **177 (44.81%)** | **Unadjusted OR (95% CI)** | **Adjusted OR (95% CI)** |
| --- | --- | --- | --- | --- |
| How long have you been on ART? |  |  |  |  |
| Less than 1 year | 30 (13.76) | 32 (18.08) | REF | REF |
| Between 1-2 years | 32 (14.68) | 20 (11.30) | 0.59 (0.28, 1.24) | 0.60 (0.28, 1.27) |
| More than 2 years | 156 (71.56) | 125 (70.62) | 0.75 (0.43, 1.30) | 0.76 (0.44, 1.34) |
| How often do you go to the health center to pick up your ART medication? |  |  |  |  |
| Monthly | 126 (57.80) | 111 (62.71) | REF | REF |
| Every 3 months | 72 (33.03) | 50 (28.25) | 0.79 (0.51, 1.23) | 0.84 (0.53, 1.34) |
| Irregularly | 20 (9.17) | 16 (9.04) | 0.91 (0.45, 1.84) | 0.92 (0.45, 1.86) |
| Adherence to ART medication |  |  |  |  |
| Complete adherence | 64 (29.36) | 36 (20.34) | REF | REF |
| Nonadherence | 154 (70.64) | 141 (79.66) | **1.63 (1.02, 2.60)** | 1.57 (0.96, 2.56) |

*Adjusted models controlling for sex and site

**Table 3: Association between specific types of mental disorder and medication adherence**

|  | **n (%)** | **Level of adherence**  **Mean (SD)** | **Any non-adherence (n=395)**  OR (95% CI) | **Level of adherence among those without complete adherence (n=295):**  Mean Diff (95% CI) |
| --- | --- | --- | --- | --- |
| Any mental condition | 177 (44.81%) | 85.03 (15.23) | 1.57 (0.96, 2.56) | **-2.99 (-5.80, -0.18)** |
| Common mental disorder | 125 (31.65%) | 85.95 (14.23) | 1.41 (0.83, 2.40) | -2.70 (-5.59, 0.18) |
| *Major depressive disorder* | 108 (27.34%) | 85.71 (14.69) | 1.37 (0.78, 2.41) | -2.62 (-5.65, 0.42) |
| *Dysthymic disorder* | 2 (0.51%) | 100 (0.00) | -- | -- |
| *Panic disorder* | 13 (3.29%) | 77.86 (26.11) | 2.07 (0.51, 8.40) | **-13.79 (-20.98, -6.60)** |
| *Generalized anxiety disorder* | 24 (6.08%) | 84.40 (12.29) | 2.52 (0.84, 7.58) | **-4.95 (-9.64, -0.27)** |
| *Post-traumatic stress disorder* | 16 (4.05%) | 88.54 (6.39) | 3.88 (0.80, 18.73) | 2.10 (-3.04, 7.24) |
| *Somatization disorders* | 3 (0.76%) | 81.11 (14.70) | -- | -- |
| Severe mental disorder | 97 (24.56%) | 83.54 (15.46) | **2.38 (1.24, 4.56)** | **-3.87 (-7.01, -0.73)** |
| *Mania* | 27 (6.84%) | 81.15 (13.73) | **4.66 (1.03, 21.17)** | -4.22 (-8.52, 0.08) |
| *Psychotic disorders* | 87 (22.03%) | 83.31 (16.07) | **2.33 (1.19, 4.56)** | **-4.39 (-7.69, -1.09)** |
| Any suicide risk (low/med/high) | 61 (15.44%) | 83.88 (17.09) | 1.28 (0.62, 2.63) | **-4.25 (-8.13, -0.37)** |
| Substance use disorder | 34 (8.61%) | 83.01 (14.69) | 2.08 (0.68, 6.38) | -3.18 (-7.32, 0.96) |
| *Alcohol use disorder* | 33 (8.35%) | 82.73 (14.83) | 2.01 (0.65, 6.18) | -3.63 (-7.83, 0.57) |
| *Drug use disorder* | 5 (1.27%) | 65.33 (23.27) | -- | -- |

Note: the reference group for each of these comparisons included participants without any mental disorders (same reference group for all comparisons). All models controlled for sex and site. The dysthymia, somatization, and drug use disorder logistic models did not converge due to small cell sizes; OR: Odds Ratio; Mean Diff: The mean difference in the level of non-adherence
